# Impact of COVID-19 on education, health and lifestyle behaviour of Brazilian urology residents

**DOI:** 10.1101/2021.01.26.21250518

**Authors:** Jose Antonio Prezotti, João Victor T. Henriques, Luciano A. Favorito, Alfredo F. Canalini, Marcos G. Machado, Thulio B. V. Brandão, Akemi M. V. Barbosa, Julyana K.M. Moromizato, Karin M. J. Anzolch, Roni de C. Fernandes, Fransber R. A. Rodrigues, Carlos H. S. Bellucci, Caroline S. Silva, Antonio Carlos L. Pompeo, Jose de Bessa, Cristiano M. Gomes

## Abstract

**Objectives:** To evaluate the impact of COVID-19 on clinical and surgical practice, educational activities, health and lifestyle behavior of Brazilian urology residents.

**Materials and Methods:** A web-based survey was sent to 468 Brazilian urology residents from postgraduate years (PGY) 3 to 5 to collect data on clinical practice and training after 4 months of COVID-19. We also assessed health-related and behavior changes, rate of infection by SARS-CoV-2, deployment to the front line of COVID-19, residents’ concerns, and access to personal protective equipment (PPE).

**Results:** Massive reductions in elective and emergency patient consultations, diagnostic procedures and surgeries were reported across the country, affecting PGY 3 to 5 alike. Most in-person educational activities were abolished. The median damage to the urological training expected for 2020 was 6.0 [3.4 -7.7], on a scale from 0 to 10, with senior residents estimating a greater damage (P< 0.001). Educational interventions developed included online case-based discussions, subspeciality conferences and lectures, and grand rounds. Most senior residents favored extending residency to compensate for training loss and most younger residents favored no additional training (p< 0.001). Modifications in health and lifestyle included weight gain (43.8%), reduced physical activity (68.6%), increased alcoholic intake (44.9%) and cigarette consumption (53.6%), worsening of sexual life (25.2%) and feelings of sadness or depression (48,2%). Almost half were summoned to work on the COVID-19 front-line and 24.4% had COVID-19. Most residents had inadequate training to deal with COVID-19 patients and most reported a shortage of PPE. Residents’ concerns included the risk of contaminating family members, being away from residency program, developing severe COVID-19 and overloading colleagues.

**Conclusions:** COVID-19 had a massive impact in Brazilian urology residents’ training, health and lifestyle behavior, which may reflect what happened in other medical specialties. Studies should confirm these findings to help developing strategies to mitigate residents’ losses.

## INTRODUCTION

COVID-19 led to profound changes in the medical scenario worldwide, including a massive reduction of face-to-face medical consultations and suspension or postponement of elective surgical procedures(1, 2). In addition, it caused a redeployment of health professionals to work at the front line caring for infected patients(3, 4). Urological practice was immensely affected. A recent survey conducted among Brazilian urologists showed an enormous reduction of patient visits, elective, and emergency surgeries, as well as a drastic reduction of income all over the country(5).

Urology residency programs have been facing problems that include not only the major cutback of residents’ participation in medical visits and surgeries, but also the reduction of educational and scientific activities (3, 4, 6, 7). Moreover, residents represent a large component of the workforce at various health facilities and in many hospitals they have become critical at the front line of care for patients with COVID-19(3, 4).

Urology residents have dealt with major challenges not only in terms of medical training, but also regarding their personal lives, health and well-being. Social distancing, fear of contamination and/or transmitting the disease to relatives and reduction of income are some of the problems and concerns affecting medical residents. Consistent with this, a significant proportion have been experiencing anxiety and depression disorders (8, 9).

While the pandemic numbers went down in many countries for some time, a second wave is effective in many regions (US, Europe)(10, 11). Some countries like Brazil, Argentina and India persist with high COVID-19 infection rates(10, 12). As a result, a significant detrimental impact on urology training has occurred and should remain indefinitely in many countries.

Considering the unique behavior of COVID-19 in Brazil, we hypothesized that Brazilian urology residents were severely impacted in their training, health and personal lives and that the impact on urology training was more severe for senior residents.

## MATERIALS AND METHODS

This study was approved by the Research Ethics Committee of the University of Sao Paulo School of Medicine (project number CAPPESQ 13029/2020) and informed consent was obtained from all participants.

An electronic survey was emailed to 468 eligible urology residents from postgraduate years (PGY) 3 to 5 registered in the Brazilian Society of Urology, with no incentives for completion. The first e-mail inviting the residents to participate was sent on June/11/2020 and data collection was closed on June/19/2020, when the whole country had lived the third month of social distance officially recommended by state and federal sanitary authorities

The primary goal was to collect data on resident’s clinical practice and urological training since the beginning of COVID-19. The survey included an assessment of socio-demographic, clinical practice, educational, health-related and behavior parameters. The invitation e-mail contained a link to a 39 questions, web-based survey (Supplementary Questionnaire). Most questions were closed-ended, multiple choice. One question evaluated the subjective degree of damage to the urological training based on the residents’ educational expectations for 2020. It was evaluated with a visual analog scale ranging from zero to ten, in which “zero” was the least possible educational damage.

We also investigated alternative educational activities that were implemented by urology residency programs such as web meetings, web conferences and online education programs.

We investigated the number of residents that were infected by SARS-CoV-2, the deployment to the front line of COVID-19 and other aspects associated with residents’ fears and concerns, and access to personal protective equipment. Changes in health parameters and behaviors such as body weight, physical activities, alcohol and tobacco consumption and sexual life were also evaluated.

Finally, based on the continental dimensions of Brazil and the varying incidence rates of COVID-19 and stages of economic opening, we compared data from states that were among the lowest incidence rates at the time of the survey (Minas Gerais, Goias, Rio Grande do Sul and Parana) with those among the highest incidence states (São Paulo and Rio de Janeiro)(12).

### Data collection and Statistical analyses

Data were initially elaborated using Survey Monkey® software online. Quantitative variables were expressed as medians and interquartile ranges, while qualitative variables were expressed as absolute values, percentages, or proportions.

Student’s t or ANOVA was used to compare continuous variables.

Categorical variables were compared using the Chi-squared or Fisher’s exact test. Associations were described as Odds Ratios with respective confidence intervals. All tests were 2-sided and a p value < 0.05 was considered statistically significant. GraphPad Prism, version 8.0.4, San Diego-CA, USA, was used for data analysis

## RESULTS

A total of 275 (58.7%) subjects completed the survey. Most respondents (90.5%) were men, and the median age was 30 years [28-31]. Ninety (32.7%) participants were from PGY 3, 90 (32.7%) from PGY 4 and 95 (34.6%) from PGY 5. The distribution of participants was proportional to the actual distribution of Brazilian urology residents across the country’s five different geographic regions (supplementary material; p=0.632).

### IMPACT OF COVID ON RESIDENTS’ CLINICAL AND SURGICAL ACTIVITIES

Major reductions in elective and emergency patient consultations, diagnostic procedures and surgeries were reported by residents from all states. Similar reductions of elective visits (p=0.398), emergency visits (0.092), minor surgeries (p=0.344), major surgeries (p=0.107), and diagnostic procedures (p=0.159) were observed across residents from PGY 3, 4 and 5. There were no differences between states with highest vs lowest incidence of COVID-19 (Table 1).

**Table 1.** Impact of COVID-19 on Brazilian urology residents’ clinical practice

| Practice activity | % |
| --- | --- |
| Elective patient visits |  |
| Remained stable | 5.9 |
| Decreased up to 25% | 8.7 |
| Decreased 25 to 50% | 20.7 |
| Decreased 50 to 75% | 28.0 |
| Decreased > 75% | 36.7 |
| Emergency patient visits |  |
| Remained stable | 31.3 |
| Decreased up to 25% | 20.0 |
| Decreased 25 to 50% | 25.8 |
| Decreased 50 to 75% | 12.4 |
| Decreased > 75% | 10.5 |
| Minor surgeries* |  |
| Remained stable | 4.4 |
| Decreased up to 25% | 9.1 |
| Decreased 25 to 50% | 8.0 |
| Decreased 50 to 75% | 13.8 |
| Decreased > 75% | 64.7 |
| Endoscopic surgeries** |  |
| Remained stable | 14.2 |
| Decreased up to 25% | 13.9 |
| Decreased 25 to 50% | 25.9 |
| Decreased 50 to 75% | 17.1 |
| Decreased > 75% | 28.9 |
| Major surgeries*** |  |
| Remained stable | 16.8 |
| Decreased up to 25% | 13.5 |
| Decreased 25 to 50% | 22.3 |
| Decreased 50 to 75% | 18.9 |
| Decreased > 75% | 28.5 |
| Diagnostic procedures |  |
| Remained stable | 9.8 |
| Decreased up to 25% | 7.6 |
| Decreased 25 to 50% | 16.7 |
| Decreased 50 to 75% | 19.6 |
| Decreased > 75% | 46.2 |
| Telemedicine implementation |  |
| Yes | 28.1 |
| No | 71.9 |

\* i.e. vasectomy, circumcision, hydrocelectomy
\*\* i.e. transurethral resection of prostate, transurethral resection of bladder tumor, ureterolithotripsy
\*\*\* i.e. oncological, laparoscopic surgeries, kidney transplantation

### IMPACT ON EDUCATIONAL ACTIVITIES

The impact of COVID-19 on urology residents’ educational program and scientific activities is shown in Table 2, with major reductions in general urology and subspecialty meetings in addition to in-person practical educational activities.

**Table 2.** Impact on educational activities during COVID-19

| **Educational and scientific activities cancelled durin COVID-19** | **%** |
| --- | --- |
| **General urology department grand rounds** | 53.8 |
| **Subspecialty conference** | 25.8 |
| **Interdisciplinary conference** | 29.1 |
| **Morbidity and mortality meeting (M&M)** | 16.0 |
| **Core Curriculum lecture** | 38.5 |
| **Journal clubs meeting** | 26.2 |
| **Bedside clinical teaching** | 54.2 |
| **Educational and scientific activities developed during COVID-19** | **%** |
| **None** | 10.9 |
| **Case based discussion webmeeting** | 68.3 |
| **Subspecialty webconference** | 40.7 |
| **Pre-recorded video lectures** | 56.0 |
| **Online courses** | 17.4 |
| **Journal clubs webmeeting** | 54.9 |
| **Online general urology department grand rounds** | 60.0 |

The median estimated damage to the urological training expected for 2020 in a scale from 0 to 10 was 6.0 [3.4-7.7]. PGY 4 and 5 residents estimated a greater damage for their training than PGY 3 (P< 0.001; figure 1). Residents living in states with highest incidence of COVID-19 did not differ from those living in lowest incidence states (p=0.777).

**FIGURE 1.**
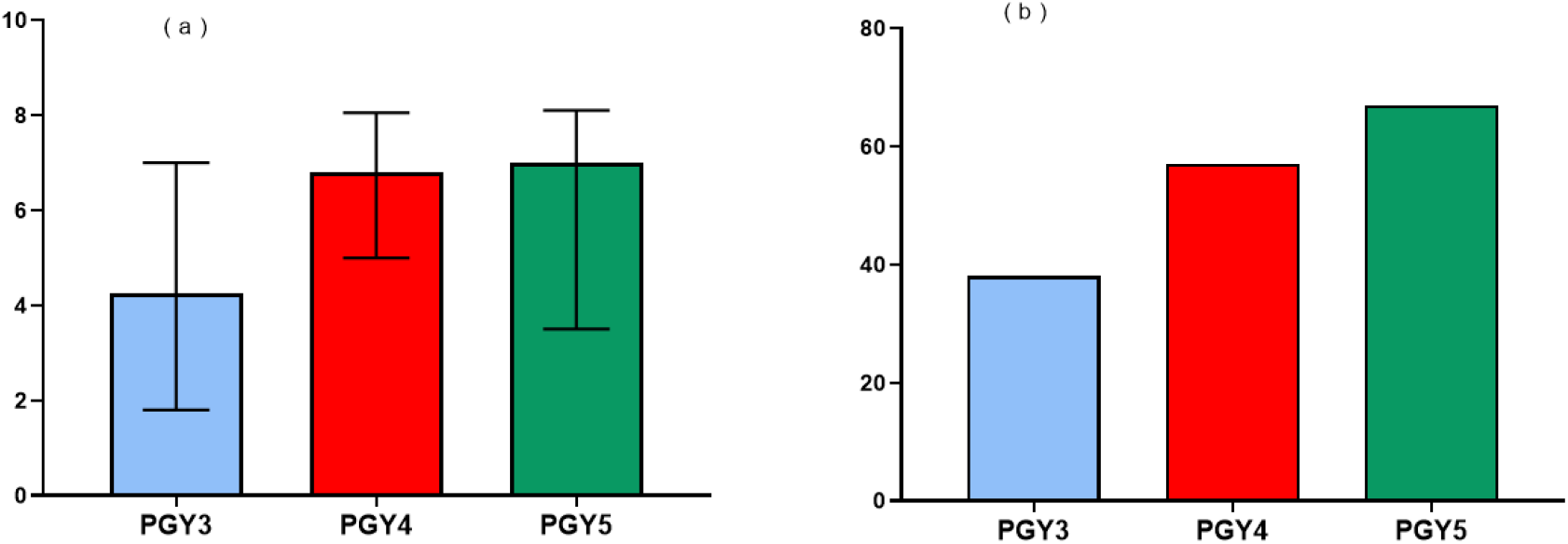
a: Damage to urological learning in 2020 as estimated by residents from postgraduate years 3, 4 and 5 b: Percentage of residents from postgraduate years 3, 4 and 5 who favor extending the residency program PGY3: Postgraduate year 3 PGY4: Postgraduate year 4 PGY5: Postgraduate year 5

### EDUCATIONAL INITIATIVES DURING THE PANDEMIC

Since the start of COVID-19 pandemic, educational interventions developed by residency programs to compensate for the deprivation of training were frequent and included: case based discussions, journal club web meetings, subspeciality web conferences, pre-recorded video lectures, online courses and online general urology department grand rounds (Table 2). Asked about their preferred educational activities, online courses and lectures were chosen as their favorite instructional method by 48.1% of the urology residents, case-based discussions web meetings by 38.2% and didactic podcasts by 14.6%.

Regarding measures that should be adopted to compensate for the loss of their urological training in 2020, 147 (53.6%) suggested postponing the finishing date of the residency program, 57 (20.8%) suggested additional training in specific subspecialties and 70 (25.6%) required no compensation. Urology residents’ perceptions regarding the need for additional training were significantly different according to postgraduate year, with senior residents favoring extending residency for some months and younger residents favoring no additional training (p< 0.001; Figure 1).

### IMPACT ON RESIDENTS’S HEALTH

The impact of COVID-19 on urology residents’ health is shown in Table 3. A total of 148 (53.8%), 54 (19.6%) and 73 (26.6%) considered themselves healthy, moderately healthy and not healthy, respectively. Weight gain was reported by 43.8% and 68.6% reported a reduction of physical activities. Of the 28 (11.4%) participants who declared being tobacco smokers, cigarette consumption increased for 15 (53.6%) and decreased for 2 (7.1%) residents. Among the participants who drink alcoholic beverages, increased consumption was reported by 44.9%. A total of 169 (61,68%), 44 (16.06%) and 61(22,26%) considered themselves satisfied, neither satisfied nor dissatisfied and dissatisfied with their sexual life, respectively. In comparison with their pre-pandemic sexual life, 163 (59.5%) subjects reported their sexual life remained stable, 69 (25.2%) reported it became worse and 42 (15.3%) reported an improvement. Feelings of sadness or depression were reported as at least sometimes by 87 (31.7%) and frequent by 45 (16.5%) residents.

**Table 3.** Health parameters during COVID-19

| Health parameters | % |
| --- | --- |
| Weight |  |
| Reduced | 14.6 |
| Stable | 41.6 |
| Increased | 43.8 |
| Physical activity |  |
| Reduced | 68.6 |
| Stable | 20.4 |
| Increased | 10.9 |
| Alcoholic beverages intake* |  |
| Reduced | 14.1 |
| Stable | 41.0 |
| Increased | 44.9 |
| Satisfaction with sexual life |  |
| Satisfied | 25.2 |
| Neither satisfied nor dissatisfied | 59.5 |
| Dissatisfied | 15.3 |
| Sexual life during COVID-19 (compared to pre-COVID-19) |  |
| Worsened | 25.2 |
| Unchanged | 59.5 |
| Improved | 15.3 |

\*participants who reported not drinking alcoholic beverages were removed from calculations

In June/2020, 67 (24.4%) of Brazilian urology residents had had COVID-19, including 18.6% with unequivocal laboratory confirmation and 5.8% with a diagnosis based on clinical, epidemiological and radiological parameters. In all instances, the clinical presentation was mild or moderate and none required hospitalization. Also, 135 (49.3%) had been summoned up to work in the front-line treatment of COVID-19 patients. Of those directly involved in the treatment of patients infected by SARS-CoV-2,70 (25.4%) reported receiving adequate training to work with such patients while 103 (37.7%) and 100 (36.6%) reported inadequate or no training, respectively.

Regarding the support received from their staff/mentors during the pandemics, 140 (51.1%) of the participants were satisfied or very satisfied, 78 (28.5%) were neither satisfied nor dissatisfied and 56 (20.4%) were dissatisfied or very dissatisfied.

Among the concerns regarding being infected with SARS-CoV-2, the risk of contaminating family members was reported by 234 (85.7%) residents, being away from the residency program and fail to complete their training by 174 (63.7%), developing COVID-19 by 129 (47.2%), being away from hospital and overloading their colleagues by 114 (41.7%) and being summoned up to work in the front line by 53 (19.4%) residents. Fifty-two (19.0%) residents changed their housing to self-isolate and avoid the risk of transmitting the disease to a family member. A shortage of PPE was reported by most residents, including N-95 masks by 124 (61.4%) residents, waterproof apron by 101 (50.8%), protective glasses or face shield by 80 (42.2%) surgical masks by 54 (28.4%) and caps by 39 (20.3%) residents.

## DISCUSSION

We have shown a massive impact of COVID-19 on urology residents’ clinical practice and education, regardless of the post graduate year or whether they train in a state among the highest or lowest incidence of COVID-19. They have all experienced the cessation or postponement of elective and preventive consultations, diagnostic procedures and non-emergency surgeries. Training was thus severely impaired and junior residents were less worried about their education breach, having evaluated the damage to their training as significantly less severe than PGY 4 and 5 residents. Moreover, significant changes in health and lifestyle were observed, with a great proportion of residents considering themselves not healthy (46.2%), reporting feelings of sadness or depression (48.2%), reporting weight gain (43.8%), reduction of physical activities (68.6%), increased alcoholic intake (44.9%) and decreased satisfaction with sexual life (25.2%). Finally, almost half of Brazilian urology residents had been summoned up to work in the front-line treatment of COVID-19 patients and 24.4% had acquired COVID-19 at the time of the survey.

Our study was performed in June/2020, when Brazil ranked second among the countries with the highest number of deaths due to COVID-19 in the world (12, 13). As we finish the preparation of this manuscript, in early December/2020, Brazil has maintained a very high incidence of COVID-19 and is experiencing a second wave as many countries worldwide(13). Governmental guidelines supporting economy closure or reopening and social distancing have varied during the year and a return to more restrictive recommendations is evolving in response to the second wave of the pandemics. As a result, a sustained detrimental impact on residents’ training is to be expected.

A total of 458 eligible Brazilian urology residents received a link to the survey, of which 275 (58.7%) participated. Because we gave no incentives to participants and we used a long questionnaire (39 questions), we consider our participation rate as very good, and in line with recent online surveys evaluating medical residents, in which participation rates varied from 45% to 60.8% (3, 4).The distribution of respondents was in accordance with the actual distribution of residents throughout the different states of the country and also regarding gender and the post graduate year, confirming a well-balanced and representative participant distribution.

We observed a greater than 50% reduction in elective patient visits, diagnostic procedures, elective surgeries and in 22.9% of urgent consultations. Residents from different PGY were similarly affected. In a study that evaluated the impact of COVID-19 in Italian residents, a severe reduction (>40%) or complete suppression (>80%) of training exposure ranged between 41.1% and 81.2% for “clinical” activities and between 44.2% and 62.1% for “surgical” activities (3). The reductions were more pronounced for residents who attended the last year of training. This possibly reflects the heterogeneity of urological training in various residency programs or different policies for restriction of surgical procedures in the two countries. Although the impact on training has been profound among all residents in Brazil, PGY 4 and 5 residents reported more severe damage to their education, which may reflect the perceived opportunity of PGY 3 urology residents to compensate for their educational losses in the following two years of residency.

Urology residency programs cancelled most educational/scientific in-person activities and developed a number of online educational programs including case based discussion and journal club web meetings, subspeciality web conference, pre-recorded video lectures, online courses and online general urology department grand rounds. (7, 14) Furthermore, local, national, and international societies have implemented various educational activities such as webinars with ample access to participants that have also contributed to mitigate residents’ educational curtailment(6, 15, 16).

Although online activities have served as a strong tool for medical education since the beginning of the pandemics, people are getting exhausted with this format(17). Being on a video call or teleconference requires more focus than face-to-face meetings and lectures because it is harder to process non-verbal cues like facial expressions, the tone and pitch of the voice, and body language(17). In addition, many of us are using video calls at work, family celebrations and interaction with friends. Everything seems to be happening in the same place, which further contributes to negative feelings for online meetings. Moreover, the online meetings remind us of the people, opportunities, and lifestyle that we have lost temporarily.

A significant impact was observed in several health indicators of urology residents. Almost half of the participants considered themselves moderately healthy (19.6%) or not healthy (26.6%). The mental afflictions resulting from the COVID pandemic may have an important role in this self-reported perception of general well-being as well as in the high rates of depressive feelings reported. Similar findings have been observed with French urology residents, of whom 56.5% reported medium to high levels of stress (8). A survey with American urology residents demonstrated that perception of access to personal protection equipment, local COVID-19 severity and perception of susceptible household members are important risk factors of mental health outcomes(18). All of these risk factors were present among Brazilian urologists.

To protect their families, many healthcare providers have chosen to isolate themselves. (19) In our study, 20.0% of urology residents changed their housing status, to avoid transmitting COVID-19 to a family member. Home confinement is also a factor that might influence alcohol misuse, physical and sexual activity (20-23). A significant decrease of physical activity has been shown in the general population during the COVID-19 pandemic(9). In the present study, 68.6% of the participants reported reduced physical activity. Many reported weight gain (43.8%) while 14.6% had weight loss. Increased alcoholic intake during the COVID-19 pandemic was reported by 44.9% of the urology residents, which was much higher than the 17.6% increase observed in the general adult Brazilian population(9). A similar finding occurred regarding tobacco consumption, which increased for 53.4% of the smoking participants while only 34.0% of the smokers in the general Brazilian population reported increased tobacco consumption during the pandemic(9). These numbers are alarming and may reflect the increased distress caused by the pandemic among health professionals.

Sexual life satisfaction is an important issue related to mental health status. A recent study with French urologists residents demonstrated that sexual intercourse is a protective factors for burnout(24). In our study, 25.2% of the urology residents reported being dissatisfied with their sexual life. Compared to pre-COVID-19 status, 25.18% of the respondents reported a worse sexual life. Similar findings showing worsening of sexual life were found among urologists in Brazil and the general population(5, 25).

A clinical infection by SARS-CoV-2 was reported by 24.4% of the participants. To our knowledge, this is the first study to report on COVID-19 infection rates among urology residents and surgical residents in general. The infection rate in Brazilian urology residents seems much higher than expected and surpass the infection rate of 13.5% reported by urologists in Brazil.(5) A recent meta-analysis showed that around 10% of healthcare personnel working in hospitals had a diagnosis of COVID-19 infection and the risk was higher among nurses.(26) In our study, none of the participants who had a clinical infection by the COVID-19 required hospitalization. This is in great contrast with the high rates of more severe COVID-9 in Brazilian urologists, among whom 34% required hospitalization.(5) A recent meta-analyses showed a prevalence of severe COVID-19 of 5% among healthcare workers, while 0.5% died because of the complications of the disease.(26) Certainly, the age difference between participants in the present study and the one with urologists in Brazil (median of 30 and 46.0 years, respectively) was the main reason for the observed difference in clinical severity of COVID-19.(27)

Involvement of residents in COVID-19 care varied substantially by specialty in different countries. Some may be caring for patients with COVID-19 during assigned rotations while others may be voluntarily redeployed to work with these patients. Half of the participants of the present study have been summoned up to work in the front-line care of patients with COVID-19. This is in contrast with urology and surgery residents in Italy, of whom only 7.7% and 14.8% were moved to the front-line care of COVID-19 patients. (3, 28)A significant proportion of participants showed preoccupations regarding being dislocated to the front-line treatment of COVID-19. Most frequent concerns were transmitting infection to family members, compromising their urological training, being infected with COVID-19 and maintenance of financial support. These concerns are in line with those reported in other studies.(28, 29)

Only 25.4% of the respondents reported receiving adequate training to work with COVID-19 patients and most residents reported insufficient availability of PPE, including surgical masks (28.4% of the participants), N-95 masks (61.4%), face shield (42.2%) and waterproof apron (50.8%). The combination of working in the front-line care of patients with COVID-19 with inadequate training for dealing with these patients and insufficient PPE availability may have contributed to the high rate of COVID-19 infection among participants. In a recent manuscript published in the New England Journal of Medicine, Gallagher and Schleyer remind us of the personal risks health care professionals face when caring for patients with communicable diseases (29). We want to join them in praising our medical students and trainees for stepping up during the Covid-19 pandemic despite their risks and fears.

Asked about what should be done to mitigate their training breach throughout 2020, most proposed extending the training program for some months. Additional training in subspecialties was also recommended by many participants. In a study conducted in India with ophthalmology residents, eighty percent felt the pandemic had damaged their medical education and suggested that extending the residency period would be necessary (30, 31)

The main strengths of this study are the large number of participants, the balanced distribution of residents from different postgraduate years and the comprehensive evaluation of various aspects of medical practice, educational activities and health and lifestyle parameters, allowing us to make a reliable picture of the impact of the pandemic on urology residents in Brazil. This is the first study to report on COVID-19 infection rates among residents of a surgical specialty. The cross-sectional design, providing an evaluation of a single snapshot in time is a limitation of the study. As the pandemics has assumed different faces throughout the year, residents’ mood and perceptions may certainly fluctuate with time. The questionnaire is long, which may make participants bored through the process of answering it. Nevertheless, the response rates were very high. Another point is that the instruments used to evaluate many of the parameters are not validated. For instance, mental health was assessed using a single question, instead of a validated questionnaire. Despite these limitations, our findings seem to present a reliable picture of the impact of COVID-19 in a number of aspects of Brazilian urology residents’ education, health and lifestyle behavior than can possibly be expanded for residents from other medical specialties. Further studies should confirm these findings and expand on the evaluation of the detrimental impacts of COVID-19 in medical residents from different specialties to help developing strategies to mitigate their losses.

## CONCLUSION

COVID-19 had a massive impact on Brazilian urology residents’ clinical practice and education. Training was severely impaired and junior residents perceived the damage to their education as less severe than senior residents. Significant changes in health and lifestyle were observed, with many residents reporting weight gain, reduced physical activity, increased alcoholic intake and cigarette consumption, worsening of sexual life and feelings of sadness or depression. Finally, almost half of Brazilian urology residents had been summoned to work on the front-line treatment of COVID-19 patients and a quarter had acquired COVID-19 at the time of the survey.

## Data Availability

The data are under the custody of the research coordinator and may be requested by the reviewers at any time.

## ACKNOWLEDGEMENTS

The authors would like to thank the participants of the study for their time. This study received no funding.

## Author contributions

**JAP:** Conception and design; acquisition of data; analysis and interpretation of data; drafting of the manuscript; critical revision of the manuscript for important intellectual content

**JVTH:** Conception and design; acquisition of data; analysis and interpretation of data; drafting of the manuscript; critical revision of the manuscript for important intellectual content

**LF:** Acquisition of data, Drafting of the manuscript, critical revision of the manuscript for important intellectual content.

**AFC:** Acquisition of data, Drafting of the manuscript, critical revision of the manuscript for important intellectual content.

**MGM:** Conception and design; acquisition of data; analysis and interpretation of data; drafting of the manuscript; critical revision of the manuscript for important intellectual content

**TBVB :** Acquisition of data, Drafting of the manuscript, critical revision of the manuscript for important intellectual content.

**AMVB:** Acquisition of data, Drafting of the manuscript, critical revision of the manuscript for important intellectual content.

**JKMM :** Acquisition of data, Drafting of the manuscript, critical revision of the manuscript for important intellectual content.

**KMJA:** Acquisition of data, Drafting of the manuscript, critical revision of the manuscript for important intellectual content.

**RF:** Acquisition of data, Drafting of the manuscript, critical revision of the manuscript for important intellectual content.

**FRAR:** Acquisition of data, Drafting of the manuscript, critical revision of the manuscript for important intellectual content.

**CHSB:** Conception and design; acquisition of data; analysis and interpretation of data; drafting of the manuscript; critical revision of the manuscript for important intellectual content

**CSS:** Acquisition of data, Drafting of the manuscript, critical revision of the manuscript for important intellectual content.

**ACLP:** Acquisition of data, Drafting of the manuscript, critical revision of the manuscript for important intellectual content.

**JBJ:** Conception and design; acquisition of data; analysis and interpretation of data; drafting of the manuscript; critical revision of the manuscript for important intellectual content

**CMG:** Conception and design; acquisition of data; analysis and interpretation of data; drafting of the manuscript; critical revision of the manuscript for important intellectual content

## Conflicts of interest

None of the authors has a financial or personal relationship that could inappropriately influence or bias the content of the paper.

**JAP:** None

**JVTH:** None

**LF:** None

**AFC:** None

**MGM:** None

**TB :** None

**AMB:** None

**JM :** None

**KMJA:** None

**RF:** None

**FRAR :** None

**CHSB:** None

**CSS:** None

**ACLP:** None

**JBJ:** None

**CMG:** Grant/Research study: Ipsen; Consultancy: Astellas Pharma, Boston Scientifics, Coloplast, Medtronic; Lectures: Astellas, Boston Scientific, Zodiac, Apsen

## Supplementary Material

Geographic regions in Brazil and state distribution of the 275 participants of the survey

| Geographic region / State | N* (%) | Actual state distribution** (%) |
| --- | --- | --- |
| Southeast | 180 (65.4) | 316 (67.3) |
| Sao Paulo | 102 (37.1) | 188 (40.1) |
| Minas Gerais | 26 (9.5) | 57 (12.1) |
| Rio de Janeiro | 48 (17.4) | 64 (13.6) |
| Espirito Santo | 4 (1.4) | 7 (1.5) |
| South | 24 (8.7) | 48 (10.1) |
| Parana | 12 (4.3) | 28 (5.9) |
| Rio Grande do Sul | 11 (4.0) | 14 (2.9) |
| Santa Catarina | 1 (0.4) | 6 (1.3) |
| Northeast | 42 (15.3) | 68 (14.4) |
| Alagoas | 0 (0.0) | 0 (0.0) |
| Bahia | 12 (4.3) | 15 (3.2) |
| Ceara | 12 (4.3) | 17 (3.5) |
| Maranhao | 0 (0.0) | 0 (0.0) |
| Paraiba | 0 (0.0) | 0 (0.0) |
| Pernambuco | 12 (4.3) | 30 (6.4) |
| Piaui | 0 (0.0) | 0 (0.0) |
| Rio Grande do Norte | 6 (2.4) | 6 (1.3) |
| Sergipe | 0 (0.0) | 0 (0.0) |
| Center-West | 24 (8.7) | 33 (7.1) |
| Federal District | 8 (2.9) | 13 (2.8) |
| Goias | 11 (4.0) | 9 (1.9) |
| Mato Grosso | 2 (0.7) | 3 (0.7) |
| Mato Grosso do Sul | 3 (1.1) | 8 (1.7) |
| North | 5 (1.8) | 6 (1.2) |
| Acre | 0 (0.0) | 0 (0.0) |
| Amapa | 0 (0.0) | 3 (0.6) |
| Amazonas | 4 (1.4) | 3 (0.6) |
| Para | 0 (0.0) | 0 (0.0) |
| Rondonia | 0 (0.0) | 0 (0.0) |
| Roraima | 0 (0.0) | 0 (0.0) |
| Tocantins | 1 (0.4) | 0 (0.0) |

\* Urology residents who participated in the survey
\*\*Official state distribution of urology residents who received invitation for the survey

## Notes

### Competing Interest Statement

The authors have declared no competing interest.

### Funding Statement

This study received no external funding.

## REFERENCES

1. Ficarra V, Novara G, Abrate A, Bartoletti R, Crestani A, De Nunzio C, et al. Urology practice during the COVID-19 pandemic. Minerva Urol Nefrol. 2020;72(3):369–75.

2. Carneiro A, Wroclawski ML, Nahar B, Soares A, Cardoso AP, Kim NJ, et al. Impact of the COVID-19 Pandemic on the Urologist’s clinical practice in Brazil: a management guideline proposal for low- and middle-income countries during the crisis period. Int Braz J Urol. 2020;46(4):501–10.

3. Amparore D, Claps F, Cacciamani GE, Esperto F, Fiori C, Liguori G, et al. Impact of the COVID-19 pandemic on urology residency training in Italy. Minerva Urol Nefrol. 2020;72(4):505–9.

4. Rosen GH, Murray KS, Greene KL, Pruthi RS, Richstone L, Mirza M. Effect of COVID-19 on Urology Residency Training: A Nationwide Survey of Program Directors by the Society of Academic Urologists. J Urol. 2020:101097JU0000000000001155.

5. Gomes CM, Favorito LA, Henriques JVT, Canalini AF, Anzolch KMJ, de Carvalho Fernandes R, et al. Impact of COVID-19 on clinical practice, income, health and lifestyle behavior of Brazilian urologists. Int Braz J Urol. 2020;46.

6. Kwon YS, Tabakin AL, Patel HV, Backstrand JR, Jang TL, Kim IY, et al. Adapting Urology Residency Training in the COVID-19 Era. Urology. 2020;141:15–9.

7. Chan EP, Stringer L, Wang PZT, Dave S, Campbell JD. The impact of COVID- 19 on Canadian urology residents. Can Urol Assoc J. 2020;14(6):E233–E6.

8. Abdessater M, Roupret M, Misrai V, Matillon X, Gondran-Tellier B, Freton L, et al. COVID19 pandemic impacts on anxiety of French urologist in training: Outcomes from a national survey. Prog Urol. 2020;30(8-9):448–55.

9. Malta DC, Szwarcwald CL, Barros MBA, Gomes CS, Machado Í E, Souza Júnior PRB, et al. The COVID-19 Pandemic and changes in adult Brazilian lifestyles: a cross-sectional study, 2020. Epidemiol Serv Saude. 2020;29(4):e2020407.

10. COVID-19 Dashboard by the Center for Systems Science and Engineering (CSSE) at Johns Hopkins University (JHU) [Internet]. 2020.

11. Coronavirus second wave: Which countries in Europe are experiencing a fresh spike in COVID-19 cases? Available at: https://www.euronews.com/2020/11/18/is-europe-having-a-covid-19-second-wave-country-by-country-breakdown. 2020.

12. Bean BP. Pharmacology and electrophysiology of ATP-activated ion channels. Trends Pharmacol Sci. 1992;13(3):87–90.

13. World Health O. Coronavirus disease (COVID-19): situation report, 162. Geneva: World Health Organization. 2020.

14. Vargo E, Ali M, Henry F, Kmetz D, Drevna D, Krishnan J, et al. Cleveland Clinic Akron General Urology Residency Program’s COVID-19 Experience. Urology. 2020;140:1–3.

15. Smigelski M, Movassaghi M, Small A. Urology Virtual Education Programs During the COVID-19 Pandemic. Curr Urol Rep. 2020;21(12):50-.

16. Campi R, Amparore D, Checcucci E, Claps F, Teoh JY, Serni S, et al. Exploring the Residents’ Perspective on Smart learning Modalities and Contents for Virtual Urology Education: Lesson Learned During the COVID-19 Pandemic. Actas Urol Esp. 2020.

17. The reason Zoom calls drain your energy: Available at: https://www.bbc.com/worklife/article/20200421-why-zoom-video-chats-are-so-exhausting. 2020.

18. Khusid JA, Weinstein CS, Becerra AZ, Kashani M, Robins DJ, Fink LE, et al. Well-being and education of urology residents during the COVID-19 pandemic: Results of an American National Survey. Int J Clin Pract. 2020;74(9):e13559.

19. Xiang YT, Yang Y, Li W, Zhang L, Zhang Q, Cheung T, et al. Timely mental health care for the 2019 novel coronavirus outbreak is urgently needed. Lancet Psychiatry. 2020;7(3):228–9.

20. Pecanha T, Goessler KF, Roschel H, Gualano B. Social isolation during the COVID-19 pandemic can increase physical inactivity and the global burden of cardiovascular disease. Am J Physiol Heart Circ Physiol. 2020;318(6):H1441–H6.

21. Rodriguez LM, Litt DM, Stewart SH. Drinking to cope with the pandemic: The unique associations of COVID-19-related perceived threat and psychological distress to drinking behaviors in American men and women. Addict Behav. 2020;110:106532.

22. Lopes GP, Vale FBC, Vieira I, da Silva Filho AL, Abuhid C, Geber S. COVID- 19 and Sexuality: Reinventing Intimacy. Arch Sex Behav. 2020.

23. Turban JL, Keuroghlian AS, Mayer KH. Sexual Health in the SARS-CoV-2 Era. Ann Intern Med. 2020;173(5):387–9.

24. Gas J, Bart S, Michel P, Peyronnet B, Bergerat S, Olivier J, et al. Prevalence of and Predictive Factors for Burnout Among French Urologists in Training. European Urology. 2019;75(4):702–3.

25. Arafat SMY, Alradie-Mohamed A, Kar SK, Sharma P, Kabir R. Does COVID- 19 pandemic affect sexual behaviour? A cross-sectional, cross-national online survey. Psychiatry Res. 2020;289:113050.

26. Gomez-Ochoa SA, Franco OH, Rojas LZ, Raguindin PF, Roa-Diaz ZM, Wyssmann BM, et al. COVID-19 in Healthcare Workers: A Living Systematic Review and Meta-analysis of Prevalence, Risk Factors, Clinical Characteristics, and Outcomes. Am J Epidemiol. 2020.

27. Romero Starke K, Petereit-Haack G, Schubert M, Kampf D, Schliebner A, Hegewald J, et al. The Age-Related Risk of Severe Outcomes Due to COVID-19 Infection: A Rapid Review, Meta-Analysis, and Meta-Regression. Int J Environ Res Public Health. 2020;17(16).

28. Pertile D, Gallo G, Barra F, Pasculli A, Batistotti P, Sparavigna M, et al. The impact of COVID-19 pandemic on surgical residency programmes in Italy: a nationwide analysis on behalf of the Italian Polyspecialistic Young Surgeons Society (SPIGC). Updates Surg. 2020;72(2):269–80.

29. Gallagher TH, Schleyer AM. “We Signed Up for This!” - Student and Trainee Responses to the Covid-19 Pandemic. N Engl J Med. 2020;382(25):e96.

30. Mishra D, Nair AG, Gandhi RA, Gogate PJ, Mathur S, Bhushan P, et al. The impact of COVID-19 related lockdown on ophthalmology training programs in India - Outcomes of a survey. Indian J Ophthalmol. 2020;68(6):999–1004.

31. Paesano N, Santomil F, Tobia I. Impact of COVID-19 Pandemic on Ibero-American Urology Residents: Perspective of American Confederation of Urology (CAU). International braz j urol. 2020;46:165–9.

